# Development and Validation of the ICIS (Inclusive Capacity and Inclusion Intent Scale): A Psychometric Scale to Measure Individual Inclusion Intent and Inclusive Capacity

**DOI:** 10.1101/2025.08.14.25333692

**Authors:** Lina María González-Ballesteros, Camila Andrea Castellanos Roncancio, Luis Eduardo Mojica Ospina

## Abstract

Violence and social exclusion are interconnected issues driven by factors such as inequality, poverty, and weak state institutions, leading to the marginalization of vulnerable groups. Being able to measure individuals’ ability to include others could be useful in guiding social intervention programs and improving resource allocation, especially in vulnerable populations. Using a mixed methods study we developed, pilot tested and evaluated a research-based scale to assess individuals’ inclusion intent as a proxy for inclusion capacity. The psychometric properties of this scale were also assessed. We found strong internal consistency for the entire scale (α = 0.89) and for two of the three domains (Attitudes α = 0.84, Perceived Control α = 0.83). Although the Subjective Norm domain showed lower reliability (α = 0.51), likely due to the limited number of items. These results provide preliminary evidence of construct validity whilst suggesting areas for enhancement, particularly regarding the measurement of the subjective norms related to inclusion. The new ICIS scale provides an innovative and reliable measure of inclusion intent, serving as an indicator of inclusion capacity. It is a useful tool for assessing individual inclusion abilities across various fields, including research, clinical practice, and policy development or implementation. Furthermore, it could be used to assess the general inclusion capacity of a community. Due to its recent introduction, direct comparisons with prior studies remain challenging. Further research is required to adjust any underperforming aspects of the scale and assess its validity in wider populations.

## 1 Introduction

Violence has become a public health problem and has been ranked as the ninth cause of death in the population between 15 and 49 years old, according to data from the Institute for Health Metrics and Evaluation (IHME). In line with this, Colombia is a country that has lived through different periods of violence (Sánchez, Bella and Díaz, 2013), a fact has generated several changes in behavior and coping patterns in adult, child and adolescent populations. Some of the factors that are present in the Colombian context (violence, common crime, and poverty) increase the exclusion and vulnerability of populations, particularly for children and adolescents.

In the same manner, social exclusion in our country is evident. This is evidenced, for example, in the fact that people with disabilities tend to have lower educational achievements, lack access to the internet, and have lower quality of both energy and health services (Montoya González and Alzate, 2023). Both violence and social exclusion are articulated phenomena that have multiple causes. These causes have been considered to involve objective conditions such as inequality, poverty, weaknesses of the state in the face of social demands, as well as subjective factors, i.e., rational individuals who use violence for their benefit and increased power (Valencia Agudelo and Cuartas Celis, 2009; Pérez-Sáinz, Calderón-Umaña and Brenes-Camacho, 2016). The consequences of violence and social exclusion, mainly the dehumanization or sub-humanization of certain demographics, result in specific groups being valued as more vulnerable and frequently seen as distinct and different, favoring their exclusion and increasing aggressiveness towards them(Haslam and Loughnan, 2014).

Although neurosciences have provided answers about brain structures related to the perceptions and categorization of individuals based on their racial, physical, and emotional differences(Singer and Lamm, 2009; Forbes and Grafman, 2010), the evidence on how this translates in terms of inclusive capacity from the perspective of the individual is scarce. The capacity to include the other as a peer has been the focus of intervention from the social sectors recently. In 2019, the American Psychological Association (APA) included the construct of inclusive capacity in its thesaurus, as the process of enhancing the equitable and comprehensive participation of individuals and groups in society(Association, 2020). However, this definition exempts it from the process that individuals exercise to include others as peers, allowing people to humanize and build peer and bidirectional relationships with others. This possibility, which is not clearly defined, occurs through processes mediated by social cognition such as empathy, theory of mind, and mirror neurons(Kwan and Fiske, 2008; Fiske, 2009; Waytz, Epley and Cacioppo, 2010).

Considering the context above, developing the necessary tools to determine the impact of social intervention programs is one of the significant challenges of organizations and institutions in Colombia. This is because impact measurements allow replication and sustainability according to their effectiveness with different populations. In general, these measurements open an array of possible adjustments ranging from the optimization of resources, based on the feedback provided by these tools, to the creation of academic scenarios where the experiences of each population can serve as an example for other scenarios(Londoño Pérez and Alejo Castañeda, 2017)

The development of a measurement tool must include a rigorous process of design, piloting, and adjustment(Boateng *et al*., 2018). Following a methodology for designing measurement instruments in health, the Saldarriaga Concha Foundation (FSC) has developed instruments to measure the effects of programs focused on the participation of people in reconciliation scenarios. Said instruments have been previously used by international and national entities in order to assess the course of actions and resource distribution. With regards to the concept of inclusion, there are various knowledge gaps worth noting, including the definition and application of the construct, measurement instruments concerning inclusion, and the assessment of biological and mental processes that motivate the attitude towards inclusion.

Thus, a research project at the FSC sought to generate a valid, evidence-based scale with a straightforward, simple application in order to measure people’s capacity for inclusion. This scale should apply to populations of diverse social contexts, educational and economic levels.

## 2 Materials and Methods

This mixed methods study aimed to develop a scale to measure people’s capacity for inclusion through their intent to include others (“inclusion intent” from now on). Researchers from Fundación Saldarriaga Concha (FSC) in Bogotá (Colombia) conducted this study.

### 2.1 Defining the construct

To define the central construct being measured (inclusion intent), we constructed a nomologic network to represent the constructs of interest and the relationships between them. This nomologic network was built based on an overview of the available literature regarding the related constructs (i.e.: diversity, equity and inclusion) via a string search generated through the PsycArticles, PsycTests and PsycBooks databases to identify the definitions or mentions of “inclusion” as a construct or “inclusion capacity” used in the literature. The definitive version of the nomologic network is included as a Figure in the supplementary material of this paper.

This nomologic network was also nourished by information gathered from a first round of focus groups conducted in eight cities from different regions of Colombia (Tumaco, Tunja, Leticia, Mitú, Manizales, Bogotá, Barranquilla, and Cali). FSC selected participants were either direct beneficiaries or people related to the foundation or partner foundations working with people with disabilities or in the community. The participants included adults from different social groups including teachers, project coordinators of interventions for inclusion in the classroom linked to FSC or to the Ministry of Education, psychologists implementing FSC interventions and members of their families, foundation managers who worked in inclusion or protection of vulnerable populations, beneficiaries of assistance programs of the Santo Domingo Foundation, graduate students of Economics. The focus groups aimed to identify ideas, perceptions, and beliefs on identifying people’s ideas, perceptions, and beliefs about inclusiveness. Each focus group was conducted in person in each of the cities. The audio of each focus group was recorded and transcribed verbally.

Subsequently, two different FSC researchers carried out a thematic analysis using two levels of analysis: descriptive/explanatory and interpretative. On one hand, the descriptive and explanatory levels of analysis revealed six families of nodes that show what the participants associate with the term inclusion, what is needed to talk about it, and what is and what differs from being inclusive. Once this first level of analysis was completed, we moved on to the interpretative component of the families of nodes. This led to the second level, in which three general concepts or categories were identified that group people’s ideas and conceptions about inclusion. The interpretative analysis yielded three general categories that grouped the ideas and concepts regarding inclusion. These categories were Social wellbeing/Dignified life, Critical thought, and Context. Social well-being and dignified life include ideas related to justice, fundamental rights, and equal opportunities as necessary for society to be inclusive. Critical thought pertains to practice based on empathy and pedagogical processes promoting social reality comprehension. Context refers to a factor that places discussions around inclusion in a multidimensional scope where the capacity to be inclusive is also determined by the context and the people who participate in it. Based on the Capacity Approach of Martha Nussbaum, this analysis allowed us to identify a series of factors promoting quality of life(Nussbaum, 2011).

### 2.2 Building the scale

After the nomologic network was established, the first version of the scale was conducted based on the drafting of the items and a second round of focus groups that were specifically guided towards the drafting of the scale items. The participants for the second round of focus groups included representatives of foundations or organizations that work with inclusion, individuals working in academic institutions, and representatives of governmental initiatives. The potential participants were contacted by call or email, and those who agree to participate were included in the focus groups.

After this second round of focus groups, we proceeded to the construction of the items that make up the scale, based on an inventory of components obtained through previous literature search and review. The item inventory was constructed based on 27 different instruments, from which some items were directly used or modified to be included in a preliminary version of the scale (Larsen, Reed and Hoffman, 1980; Davis, 1983; Pettigrew and Meertens, 1995; Steenbergen, 1996; Wahl, 1999; Björkman, Svensson and Lundberg, 2007; López-Pérez, Fernández-Pinto and Abad García, 2008; Spector, Bauer and Fox, 2010; Rojas Tejada *et al*., 2011; Morselli and Passini, 2012; Reysen and Katzarska-Miller, 2013; Campo-Arias, Oviedo and Herazo, 2014; Pérez-Sáinz, Calderón-Umaña and Brenes-Camacho, 2016; Passini and Morselli, 2017; Yildirm and Türkglu, 2017; Klos and Lemos, 2018; Carrillo Sierra *et al*., 2018; Kenny, Bizumic and Griffiths, 2018; López Tello and Moreno Coutiño, 2019; Pérez-Fuentes *et al*., 2019; Tengelin *et al*., 2019; Chin *et al*., 2020; Janssen *et al*., 2020; Steindl *et al*., 2021; Arcieri and DeLucia, 2022; Martínez-Ramos, Cárdenas and Aguirre-Acevedo, 2022; Etchezahar *et al*., 2022; de Freitas Nery *et al*., 2023). The identified instruments were then sent to an expert panel with each item that made up the scale. The objective of this round was to identify the items considered candidates to integrate the preliminary version. This preliminary version grouped items within three domains: Attitudes, Perceived Behavioral Control, and Subjective Norm.

After the preliminary scale version was developed, we conducted a formal expert consensus technique to reduce the number of items in the inclusion intent scale, using the Nominal Group Technique(Chapple and Murphy, 1996). The method has four fundamental steps: 1) the individual (silent) generation of ideas, opinions, or responses; 2) the review of peer responses with the opportunity to adjust, modify, or alter their responses; 3) the clarification and clarification of opinions or concepts; and 4) voting. A web-based application was used to allow the experts to conduct the two initial phases asynchronously and remotely, which ensured that several of the key characteristics of a consensus method were met (anonymity, which avoids dominance; interaction, which allows participants to change their own opinions or responses; and controlled feedback, which gives participants access to the information provided by the group). The expert panel included a total of 11 individuals with 100% participation in each of the stages of the process. These experts were recruited through convenient sampling considering their occupational experience with inclusion and the ones who agreed to participate were included. A more detailed description of the experts can be found in the supplementary material for this article.

During the consensus, the scale items were grouped into nine dimensions (openness to change, citizenship, compassion, critical awareness of the norm, diversity, empathy, inclusion fatigue, humanitarianism, and interpersonal reactivity). During the first round (individual generation), the experts were offered the original item bank grouped by dimensions. They were asked to order them from the most to least relevant according to each dimension. They were also offered the space to submit new items or modified versions of the original ones. During the second round, the experts were provided with a stacked bar chart showing the number of votes each item had received in each possible position, the list of the original items, and those submitted by the panel members so that they could review and adjust the order generated in the previous phase. The results of this phase were analyzed to generate a coefficient reflecting the relevance of each item for the total number of participants based on the number of votes received by each item relative to the position. Based on this coefficient, the items that accumulated up to 85% relevance were selected.

During the third round, the experts were provided with the items selected based on the relevance coefficient for their final ranking. Finally, two synchronous, remote clarification and voting meetings were held in which the experts voted on the permanence within the scale of each item. Only those items that reached 70% agreement were retained. For most dimensions, an initial vote was taken, and participants had the opportunity to justify or question the items. This process included, on some occasions, adjustments in the wording of the items, the creation of new items, or the elimination of redundant items. After the clarification phase, a new vote was taken, maintaining the rule of item conservation. For some dimensions, the experts discussed the content of the items before voting. Only when a point of agreement with the content and formulation of the items was reached was the vote taken.

After the expert consensus, the generated version of the scale was subjected to a cognitive interview process with people from the scale’s target population, i.e., the general population, to assess the items’ comprehension, readability, and emotional charge. Sixteen cognitive interviews were conducted following a selective proxy purposive sampling, which means that subjects will be selected to have participants represent a combination of the relevant attributes (life course, gender, educational level, and presence of disability). This process aimed at improving the quality of the scale and to identify possible measurement problems that may arise during the response process so that the necessary adjustments can be made.

The analysis strategy for the interviews involved familiarization with the interviews by all interviewers. The interviewing team listened to all the interviews and contributed comments, suggestions, and solutions to identified problems. Two meetings were held to fine-tune the scale, in which the findings were discussed, and decisions were made on the modification of instructions, items, and response options, as well as the elimination of items. The categories and definitions used for this process were taken from those proposed by Smith-Castro and collaborators(Smith-Castro and Molina Delgado, 2011).

### 2.3 Pilot trial

After adjusting the scale items based on the cognitive interviews, the latest version was tested the first time using the JotForm platform (Inc, 2025) and distributed using a “voice-to-voice” strategy (convenience sampling) through social networks and social media of friends and family members. Forms were collected from 59 individuals (the estimated initial pilot size was 50). These forms were subjected to a descriptive analysis and discrimination indexes per each domain, to identify the items that did not adequately discriminate between the people with high and low levels of inclusion (items that present little variability and a strong tendency towards one response option).

Regarding the discrimination indexes, high positive values (tending to 1) indicate that many more subjects in the higher group answered the item with the higher inclusive ability indicator compared to the lower group, meaning the item discriminates well. Negative values indicate that more subjects from the lower group answered the item with the higher inclusive ability indicator, which is undesirable. Values close to 0 indicate that the item does not discriminate well between the groups since both groups performed similarly on the item. Items that yielded a low or marginally moderate discrimination indexes that did not affect the internal consistency of the domains were eliminated.

To preliminarily evaluate the internal consistency of the scale, a Cronbach’s α coefficient was calculated for each item to assess how well the scale’s items were related to each other and whether they measured the same concept or construct, both for the complete scale and for each domain. This analysis aided in the elimination of items that should be eliminated without hindering the internal validity of the domains.

### 2.4 Evaluation, validation, and analysis of psychometric properties of the scale

Two data collections were carried out to evaluate the psychometric properties. Regarding the first data collection, individuals who were involved with the FSC were invited to complete the scale on two different occasions, separated by a maximum of 15 calendar days. This group consisted of 37 individuals.

On the other hand, participants in the 3C strategy administered the scale before and after receiving the modules that make up the strategy. The 3C strategy is a program created by the FSC in 2015 to provide psychosocial welfare to the community of caregivers of children in prolonged crisis, because of the difficulties of internal displacement and armed conflict that the country has experienced. The strategy has evolved and is implemented to strengthen peoplés resilience and compassion. This group consisted of 400 people. The evaluation of the psychometric properties included the data from the 400 participants in the 3C strategy and was carried out as follows:

**I. Evidence of convergent validity:** The scale was applied together with the ECOM scale proposed for the measurement of compassion. It was assumed that the two constructs, inclusion and compassion, are related but measure different attributes; therefore, mild to moderate correlation coefficients were expected.
**II. Evidence of discriminating validity:** The scale was applied together with a question on perceived inclusion, and correlations were made between the full scale and the subscales with this question.
**III. Evidence internal structure validity:** With the first application of the scale, based on the proposed structure of dimensions, a confirmatory factor analysis model was constructed. The responses to each of the items were used for this purpose.
**IV. Evaluation of the internal consistency:** This was conducted with the first application of the scale, using the responses to each item.
**V. Effect size:** After a first application of the scale, participants received the 3C strategy intervention aimed at strengthening resilience and compassion. A second application of the scale followed this. Given that these constructs are related to inclusive capacity, a moderate effect on the scale’s scores (total score of the scale or in each of its dimensions) was expected.

### 2.5 Percentile transformation and reference intervals

In order to facilitate the score interpretation obtained by the participants in the validation study, a transformation process was carried out. This did not alter the scaling of the direct scores. In other words, the relative position of each individual according to the value of their scores was preserved. Moreover, this preservation of the initial order allows the scores to acquire a mainly practical utility.

We used a transformation attributed to Hazen(Hazen, 1914), which calculates an index using: *0.5 + pn.* This equation was implemented in the MedCalc statistical package as a single method for estimating reference intervals(‘MedCalc® Statistical Software version 23.2.1’, 2025). Reference intervals were further estimated with the percentile method. In this method, the lower and upper limits of normality are given by the 2.5 and 97.5 percentiles for a bilateral reference interval. Following Clinical and Laboratory Standards Institute guidelines, 95% confidence intervals are defined using the method of Reed et al(Reed, Henry and Mason, 1971). Since the distribution was skewed, the process was carried out using a logarithmic transformation, following the recommendations of Schoonjans et al(Schoonjans, De Bacquer and Schmid, 2011). The reference intervals were calculated using a nonparametric percentile method, and 95% confidence intervals were estimated for the reference limits.

The sociodemographic and clinical variables were described through central tendency and dispersion measures and using absolute and relative frequencies. Convergent validity was evaluated using the Spearman correlation coefficients, given that the data did not follow a normal distribution. The discrimination analysis of each of the subscales was carried out by means of polyscale correlation coefficients. The internal structure of the scale was evaluated by means of confirmatory factor analysis, for which a model consisting of three latent variables (attitudes, subjective norm, and perceived control) was constructed, and a correlation was assumed between them. Path coefficients, variances, and covariances were estimated using the diagonalized weighted least squares method, considering that the observed variables were ordinal. Then, the fit of this model was evaluated employing the Chi-square/degrees of freedom, RMSEA, SRMR, CFI, TLI, NFI, NNFI and IFI statistics. Three items with standardized coefficients equal to or less than 0.20 were eliminated. The lavaan library was used to build the model.

The analysis of the internal consistency of the scale and each of its subscales was carried out by means of Cronbach’s Alpha and McDonald’s Omega coefficients. The change in the coefficient was analyzed by removing the items of each subscale one by one.

The assessment of the effect size was performed by repeated measures analysis of variance, and δ Cohen’s effect size was calculated. The analysis of the minimum detectable change was performed by considering the standard deviation and the intraclass correlation coefficient (ICC) of each domain. The analyses were performed in the statistical package Stata 18 ® and in the R programming language using the R Studio environment (Team, 2022; StataCorp, 2023).

### 2.6 Ethical Aspects

The research protocol was approved by the Ethics Committee of Dexa Diab. The participation in any part of the study was free and voluntary. All participants of the study underwent an informed consent process. The authors used generative artificial intelligence tools, specifically Grammarly application for grammatical and spelling checks, as well as to suggest improvements in the writing during the preparation of the manuscript. All AI-generated suggestions were critically evaluated and modified at the authors’ discretion to ensure accuracy, scientific integrity and accountability of the final content.

## 3 Results

### 3.1 Definition of the construct

#### 3.1.1 Descriptive analysis of the focus groups

The data gathered from the first focus groups aimed at defining the construct “inclusion intent” yielded six different families of nodes: definition, perceptions, actions to improve, inclusive person, excluding person, and a person who is different.

**Definition of inclusion** refers to how the participants explained the concept of inclusion by identifying its characteristics, limits, etc. Initially, inclusion was perceived as something related to openness that derives from recognizing that we are all different. However, the concept evolved as the focus group progressed as a force that yields a life transformation for each person depending on their abilities and quality of life. Achieving the above would entail responding to what each person needs to develop fully in society, so participation in the contexts they are immersed in makes sense. In short, inclusion is a call to be part of a whole in which diversity is the key to learning to live together.

The **perception** of inclusion concerns how the participants view reality (occurrences, objects, change) for inclusion. In other words, their interpretation of external or internal information generates a mental image fed by experience and individual needs. According to the participants, inclusion is both an individual and a social issue, where stereotypes or labels only have a place when we make them explicit, distancing the other from the naturalness that makes us different. Therefore, having a disability should not be synonymous with exclusion but a call for empathy. Now, other participants also highlighted that inclusion cannot be separated from the purpose of welfare and social justice in a country like Colombia, also understanding that it is a concept that has been transformed due to the conditions of the country and global demands.

Regarding the **actions to improve inclusion** the participants mentioned that the institutional barriers that exist must disappear for institutions to become inclusive. These actions include structural actions that could improve academic processes in institutions and cultural changes, given that culture and the willingness to change were perceived to be key in the transformation process for certain practices.

An **inclusive person** alludes to the characteristics of a person who is inclusive and promotes inclusion as a social practice to promote the communitýs wellbeing that the participants identified. In summary, the most prominent characteristics related to leadership skills and bravery to face the social and ethical barriers that appear in daily life with determination.

An **excluding person** alludes to the characteristics of someone who excludes and promotes scenarios of discrimination and segregation in their daily life. It also alludes to actions that may happen unconsciously. The participants highlighted lack of empathy, the use of peoplés differences to instigate segregation, and the impossibility of dialogue as some key characteristics of these types of individuals.

A **person who is different** responds to the personification and traits that the participants used to describe someone who is unlike them or the complete opposite. Some participants used words or short phrases that alluded to gender, race, sexual orientation, type of work or dress, among other characteristics. However, most participants mentioned traits such as rude, judgmental and not listening to others.

#### 3.1.2 Interpretative analysis of the focus groups

##### Social wellbeing

One of the main ideas discussed in the focus groups was the perception of social wellbeing as a condition that hoards different factors that promote quality of life through meeting certain fundamental rights, the access to opportunities and justice. Colombia, as a Social State of Law, is obliged to guarantee fundamental rights to promote prosperity and general wellness of the nation.

As a component of social well-being, participants recognized justice as an essential ingredient, considering Colombiás history of violence and internal armed conflict. Some participants mentioned that inclusion is thus a result of justice. Moreover, it was also considered a necessary tool for the quality of a dignified life that must not only be at the hands of the State, who must guarantee the access to these rights, but also the need for action from some key individuals, such as social leaders, in promoting inclusion.

As another component of social well-being, the concept of equality of opportunities is linked to the rights approach as another line of action that the institutional framework must provide. Given that conditions of inequality and vulnerability exist, access to certain rights is obstructed. In this sense, the role of justice is to reduce existing barriers in terms of rights and offer more and better opportunities. Access to education, better working conditions, and recreational spaces, among others, are essential for inclusion to have a space for realization. Participants also mentioned that despite the differences in a country as diverse as Colombia, more opportunities would allow for more equitable scenarios, particularly in the work field.

##### Critical thought

About this category, it is important to mention that the capacity of critical thought, understood as the ability to make judgements related to what is right and to reason critically, we include notions regarding empathy, understanding diversity and openness to differences.

Regarding empathy, participants explored the concept as like to affect and the ability to identify otheŕs feelings, which allows a person to reach a consensus despite differences. Some participants also relate empathy to the lack of opportunities that people with disabilities, for example, could face. A participant mentioned the importance of recognizing our possibility of error as humans, our capacity for conscious or unconscious harm towards others, and the derived shame.

One of the participants brought up the concept of “reasonable adjustments” defined as the adequate and necessary adaptations or modifications to guarantee that anyone can benefit from their rights on equal terms. For example, access ramps were mentioned in buildings for people with reduced mobility. These adjustments are not limited to infrastructural changes but would aim to include changes in scheduling, curriculum, activities, or any material that can facilitate access for all types of people.

Regarding pedagogical processes, participants mentioned that self-criticism as an exercise within educational institutions is key to promote inclusion, given that within these institutions there is an openness towards diversity and to building and understanding reality from different points of view. There were also mentions regarding the need for deconstruction and reformulation of concepts within educational institutions that still have discriminating practices.

##### Context

Focusing on the context and environment aspects of inclusion allowed the participants to identify inclusion as a spectrum (inclusion/exclusion) in which different levels depend on the space-time context and the participants that make up this context.

On the other hand, participants also differentiated the concept of integration, defined as the comprehension of diversity and singularity of different individuals for them to live in equity, from that of inclusion, given that the former implies a difference already recognized in a group of individuals. There were mentions of other concepts such as exclusion, in the sense that it is a counterpart of inclusion that could help give perspective to some situations; segregation and vulnerability are factors directly linked to the lack of opportunities, freedom, and freedom of speech.

In full, the ideas that emerged from the focus groups on the concept of inclusion are varied and very focused on the individual perspective of what each person can do to achieve an environment where fundamental rights are respected. In turn, empathy and the concept that we are all part of the same society make the patterns of association of premises and values always leave in first place the identification of the other as an entity that requires a world in which equity prevails to achieve a dignified life.

### 3.2 Building the scale

#### 3.2.1 Expert consensus

As mentioned in the methodology section, the preliminary drafting of the scale was generated based on the items of 27 existing instruments. The items used or modified, and their instrument of origin may be found in the supplementary material of this paper. Regarding the expert consensus for the preliminary drafting of the instrument, Table 1 shows the number of items offered in each round and selected clarification and voting by domain.

**Table 1:** Results of the expert consensus rounds and number of final items.

| Domain | Number of items offered |  |  | Number of final items |
| --- | --- | --- | --- | --- |
|  | Round 1 | Round 2 | Round 3 |  |
| Openness to change | 5 | 12 | 6 | 4 |
| Citizenship | 11 | 19 | 9 | 5 |
| Compassion | 12 | 16 | 7 | 4 |
| Critical consciousness of the norm | 3 | 6 | 4 | 4* |
| Diversity | 13 | 20 | 9 | 5 |
| Empathy | 9 | 11 | 6 | 5 |
| Inclusion fatigue | 5 | 6 | 4 | 3 |
| Humanitarianism | 5 | 7 | 5 | 3 |
| Interpersonal reactivity | 5 | 9 | 4 | 3 |
| **TOTAL** | 68 | 106 | 54 | 32 |
| *\*During the meeting, one of the items was replaced by another* |  |  |  |  |

#### 3.2.2 Cognitive interviews

Sixteen (n =16) cognitive interviews were conducted according to the pre-specified sampling, seeking to reflect moments in the life course, educational level, and sex. Half of the participants were adults (18-59 years old), and the other half were 60 or older. Half of the participants were men, and half were women. Half of the participants have a complete or incomplete primary/secondary level education, whereas the other half have a complete or incomplete university education. Half of the participants had a disability.

Two instrument adjustment meetings were held in which the findings were discussed based on categories and definitions proposed by Smith-Castro and collaborators. These categories described situations or problems regarding the items themselves according to what was found in the interviews. The categories were: vagueness or contradiction, difficult technical terms (comprehension), inadequate or illogical assumptions, double barrel (two attitudinal objects in one sentence), lack of knowledge, unformed attitude, complex mental process, sensitive content, social desirability, confusing answers, inadequate answers, inconsistency between the item and its answer categories, and response overload. The research team added this last category. Based on the discussion and the analysis, items were either modified or eliminated.

#### 3.2.3 Pilot trial

After the modifications done in the previous step, the scale was distributed and applied to 59 participants. Table 2 shows the sociodemographic characteristics of the participants of the pilot trial.

**Table 2:** Sociodemographic characteristics of the pilot trial participants.

| **Sociodemographic Characteristics** |  |
| --- | --- |
| **Age, years\*** | 37.8 (15.5) |
| **Sex** |  |
| Male | 22 (37.3 %) |
| Female | 37 (62.7 %) |
| **Sexual Orientation** |  |
| Heterosexual | 53 (89.8 %) |
| Homosexual | 3 (5.1 %) |
| Bisexual | 2 (3.4 %) |
| Other | 1 (1.7 %) |
| **Gender** |  |
| Male | 22 (37.3 %) |
| Female | 36 (61 %) |
| Trans women | 1 (1.7 %) |
| **Ethnicity** |  |
| None | 55 (93.2 %) |
| Afro | 2 (3.4 %) |
| Other | 2 (3.4 %) |
| **Disability** |  |
| None | 47 (79.7 %) |
| Visual | 7 (11.9 %) |
| Mental or psychosocial | 1 (1.7 %) |
| Other | 4 (6.8 %) |

The mean age of the participants for the pilot trial was 37.8 years (SD = 15.5), and 62.7% (n = 37) of the participants were female. In addition to this, most of the participants reported being heterosexual and reported no ethnicity. Only 20.3% (n = 12) of the participants in the pilot had some kind of disability.

Regarding the internal consistency of the items and domains, the calculated Cronbach’s α for the entire instrument and each of the domains are shown in Table 3.

**Table 3:**
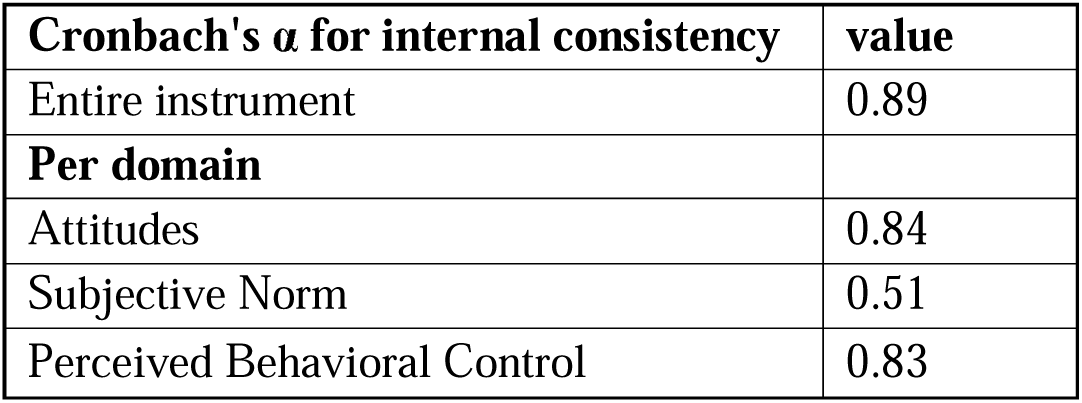
Internal consistency results for the scale.

Considering the discrimination indexes and reliability data generated during this phase of the study, a total of eight items were eliminated from the scale. The eliminated items were non-discriminatory or moderately discriminatory, and their elimination did not affect the reliability of the whole domain from which they were eliminated.

### 3.3 Evaluation, validation, and analysis of psychometric properties of the scale

A total of 438 participant responses were included. Table 4 shows the sociodemographic characteristics of the participants of this phase of the study. For the 438 participants, the median age was 29 years (IQR: 22-41), and 81.3% corresponded to the female sex. A total of 10.4% of the participants identified themselves as indigenous, and 6.8% reported having a disability.

**Table 4:** Sociodemographic characteristics of the scale evaluation participants.

| **Sociodemographic Characteristics** |  |
| --- | --- |
| **Age, years\*** | 37.8 (15.5) |
| **Sex** |  |
| Male | 82 (18.7 %) |
| Female | 356 (81.3 %) |
| **Nationality** |  |
| Colombian | 429 (98.4 %) |
| Other | 7 (1.6 %) |
| **Ethnicity** |  |
| Indigenous | 45 (10.4 %) |
| Afro | 31 (7.1 %) |
| Palenque | 1 (0.2 %) |
| Raizal | 1 (0.2 %) |
| Other | 1 (0.2 %) |
| None | 357 (81.9 %) |
| **Disability** |  |
| Physical | 9 (2.1 %) |
| Sensory | 5 (1.1 %) |
| Cognitive | 2 (0.4 %) |
| Other | 14 (3.2 %) |
| None | 408 (93.2 %) |
| *\*Median (Interquartile Range)* |  |

The description of the items that make up the three subdomains of the instrument can be found in the supplementary material of this article. For the “attitudes” domain, we found that most participants reported being in complete agreement with all the items, except for the item “I am tired of hearing about diversity-related issues,” where half of the participants reported being in complete disagreement. Regarding the “Norm” domain, we found that more than 50% of participants tend to answer “always” for all items. Finally, for the “Control” domain, most participants report that interacting with and tolerating people different from them is easy and important. On the other hand, it is reported that for most participants, it is difficult and essential to reject people who are different from them.

Table 5 presents the numerical scores obtained for each domain and the entire scale. They are reported in medians due to the non-normal distribution of the data.

**Table 5:** Numerical score results for each domain yielded in the scale evaluation.

| **Domain** | **Median Score (IQR)** |
| --- | --- |
| Entire instrument | 186 (171-201) |
| Attitudes | 96 (87-104) |
| Subjective Norm | 13 (10-14) |
| Perceived Behavioral Control | 80 (72-85) |

#### Convergent validity assessment

Slight to moderate correlations were found between our inclusion intent scale and the ECOM; the subscale of compassion towards animals was not included in the analysis. Table 6 presents the domain-to-domain correlations and the entire scale correlation. Given that the measures correspond to two related constructs, the correlations are sufficient to provide evidence of convergent validity.

**Table 6:** Domain to domain correlations and entire scale correlations (Convergent validity results)

| **Inclusion Scale Domain** | **Compassion Scale** | **Correlation** |
| --- | --- | --- |
| Attitudes | Motivation to relieve suffering | 0.4197* |
| Subjective Norm | Motivation to relieve suffering | 0.2911* |
| Perceived Behavioral Control | Motivation to relieve suffering | 0.0630 |
| **Entire scale** | Motivation to relieve suffering | 0.3308* |
| Attitudes | Affects reaction to suffering | 0.3233* |
| Subjective Norm | Affects reaction to suffering | 0.1368* |
| Perceived Behavioral Control | Affects reaction to suffering | -0.0474 |
| **Entire scale** | **Affects** reaction to suffering | 0.2001* |
| Attitudes | ECOM | 0.4178* |
| Subjective Norm | ECOM | 0.2665* |
| Perceived Behavioral Control | ECOM | 0.0264 |
| **Entire scale** | ECOM | 0.3111* |
| **\*Statistically significant correlations** |  |  |

#### Evaluation of the effect size

Table 7 presents the results of the intervention effect size.

**Table 7:** Effect size results before and after the 3C intervention.

| Scale | Score before 3C | Score after 3C | F (gl) | p-value | Effect size |  |
| --- | --- | --- | --- | --- | --- | --- |
| | Mean (SD) | Mean (SD) | | | $\delta$ Cohen | CI 95 % |
| **Attitudes** | 92.59<br>(16.19) | 91.13<br>(16.11) | 2.92<br>(1,401) | 0.0882 | 0.0903 | (-0.0379; 0.2186) |
| **Subjective Norm** | 12.06<br>(2.52) | 11.90<br>(2.43) | 1.63<br>(1,401) | 0.2027 | 0.0652 | (-0.0876; 0.1826) |
| **Perceived Behavioral Control** | 79.97<br>(13.65) | 80.19<br>(13.37) | 2.30<br>(1,401) | 0.1301 | -<br>0.0905 | (-0.2184; 0.0668) |
| **Total Score** | 184.61<br>(24.50) | 184.22<br>(24.09) | 0.09<br>(1,401) | 0.7654 | 0.0164 | (-0.0982; 0.1574) |

#### Assessment of the internal structure of the scale

We developed a structural model to create the scale based off the theoretical model proposed in previous phases. This model evaluates the relationship between the latent and observed variables. The model was adjusted to the theoretical model proposed for the development of the scale. The structural model of the scale is presented as a Figure in the supplementary material of this paper.

The removal of three items from the attitudes subdomain that had factorial loads less than 0.20 allowed us to obtain better fit statistics. Table 8 details the coefficients of the reduced model.

**Table 8:** Coefficients of the reduced model.

| $\chi^2$ | GI | $\chi^2/df$ | RMSEA | SRMR | CFI | TLI | NFI | NNFI |
| --- | --- | --- | --- | --- | --- | --- | --- | --- |
| 6.658.481 | 626 | 10.64 | 0.155 | 0.118 | 0.916 | 0.910 | 0.908 | 0.910 |

#### Internal consistency assessment

The internal consistency for the entire scale and each domain was adequate and acceptable, except for the omega coefficient for the Perceived Behavioral Control. Table 9 presents the internal consistency coefficients for each domain and the entire instrument.

**Table 9:** Internal consistency coefficients of the scale.

| Domain | $\alpha$ Coefficient | $\Omega$ Coefficient |
| --- | --- | --- |
| Attitudes | 0.933 | 0.948 |
| Subjective Norm | 0.874 | 0.824 |
| Perceived Behavioral Control | 0.672 | 0.125 |
| Entire scale | 0.833 | 0.710 |

In the supplementary material, we present the coefficients for when each item is eliminated for the subdomains of attitudes and perceived control. Since the subdomain of subjective norm has only two items, this analysis was not performed for this subdomain.

#### Discriminant validity assessment

Table 10 presents the correlation coefficients between the inclusion self-report question, the subdomains, and the entire scale. An acceptable discriminant capacity was found for the subdomains and the entire scale.

**Table 10:**
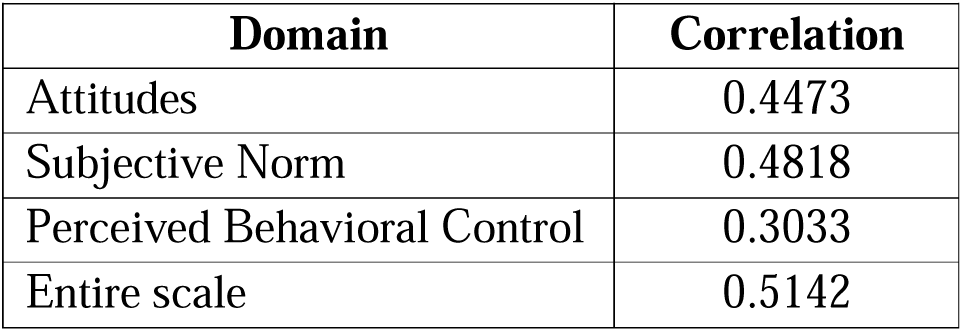
Correlation coefficients for the inclusion self-report question, the subdomains and the entire scale (Discriminant validity)

| Domain | Correlation |
| --- | --- |
| Attitudes | 0.4473 |
| Subjective Norm | 0.4818 |
| Perceived Behavioral Control | 0.3033 |
| Entire scale | 0.5142 |

#### Assessment of the minimum detectable change

Table 11 presents the results of the analysis of the minimum detectable change for each of the subdomains and the total score. This value represents the observable change in scores that must occur between two measurements for the variation to be beyond chance.

**Table 11:** Results of the minimum detectable change analysis.

| Domain | SD | ICC | MDC |
| --- | --- | --- | --- |
| Attitudes | 16.9989 | 0.6315 | 29 |
| Subjective Norm | 2.7017 | 0.3016 | 6 |
| Perceived Behavioral Control | 8.2582 | 0.3754 | 10 |
| Entire scale | 19.7302 | 0.4839 | 39 |
| **SD:** Standard Deviation; **ICC:** Intraclass Correlation Coefficient; **MDC:** Minimum Detectable Change |  |  |  |

#### Percentile transformation and reference intervals

Table 12 shows the score transformation for percentiles and their corresponding 95% confidence intervals for the ICIS scale. The score transformation for percentiles divided by domains is found in the supplementary material for this paper.

**Table 12:** Percentiles for the total scale of the ICIS scale.

| Percentile<br>s | Score | 95% CI |
| --- | --- | --- |
| **0.1** |  |  |
| **0.25** | 55 |  |
| **0.5** | 86.7359 |  |
| **1** | 111.0992 | 55.0000 - 125.6374 |
| **2.5** | 129.8746 | 112.6994 - 138.8102 |
| **5** | 145.2494 | 134.0000 - 148.0000 |
| **10** | 151 | 148.0000 - 154.8997 |
| **20** | 161 | 159.0000 - 163.0000 |
| **25** | 165 | 162.0000 - 167.0000 |
| **40** | 174 | 170.0000 - 177.0000 |
| **60** | 185 | 183.0000 - 188.0000 |
| **75** | 194 | 192.0000 - 196.0000 |
| **80** | 197 | 194.0000 - 199.0000 |
| **90** | 204 | 201.0000 - 207.0000 |
| **95** | 213.246 | 207.2414 - 218.0000 |
| **97.5** | 219.1248 | 216.1889 - 227.4450 |
| **99** | 229.8491 | 222.3875 - 248.6753 |
| **99.5** | 241 |  |
| **99.75** | 245.9839 |  |
| **99.9** |  |  |

## 4 Discussion

The construction and validation of the ICIS scale poses a relevant contribution to the social and health sciences field, particularly in contexts affected by violence and structural exclusion, such as Colombia. This study has produced an empirically grounded and psychometrically sound scale to measure the construct of inclusion intent as a proxy of inclusion capacity. This construct encapsulates an individual’s willingness and perceived ability to include others, particularly those perceived as different or ostracized.

Regarding the psychometric analysis, we found strong internal consistency for the entire scale (_α_ = 0.89) and for two of the three domains (Attitudes _α_ = 0.84, Perceived Control _α_ = 0.83). Although the Subjective Norm domain showed lower reliability (_α_ = 0.51), this finding is likely due to the limited number of items. The confirmatory factor analysis supports the proposed structure of the scale and model fit improved after the removal of the underperforming items. These results provide preliminary evidence of construct validity whilst still suggesting areas for enhancement, particularly regarding the measurement of the subjective norms related to inclusion.

Regarding the evidence of convergent validity evaluated with the ECOM scale, specifically its subscales related to compassion and affective responses to suffering, we find that the relatively modest correlations indicate that inclusion intent captures a distinct, though discretely related, psychological construct. On the other hand, the observed correlations with self-reported inclusion perceptions support the discriminant validity of the instrument.

With regards to the internal structure of the scale, it is important to mention that the RMSEA and χ^²^/gl coefficients reflect a questionable fit, despite the incremental coefficients being acceptable. This result may be due to the complexity of the model, the ordinal structure of the data or the presence of items with low factor loadings. In any case, some of the adjustment coefficients are adequate and some are insufficient, which could warrant further simplifications and adjustments in future validation and improvement processes.

The application of the scale within the context of the 3C intervention allowed for an assessment of the effect size. Considering that the 3C strategy focuses primarily on resilience and compassion, it is not surprising that the effect size is marginal regarding inclusion intent, as the intervention mainly affects aspects with a tangential relationship with the evaluated construct. Interventions focused on inclusion may be required to generate a minimally acceptable effect (of at least 0.3, as suggested by some authors).

### Comparison with Literature

Given the novelty of the ICIS scale, the possibility of conducting a direct comparison with previous studies is limited. However, it is possible to discuss how the concepts it assesses relate to theories and findings in the fields of social psychology and inclusive education. This connection helps to validate the theoretical approach of the scale and its relevance in future studies.

### Strengths

The ICIS scale was specifically developed for this study, filling a gap in the existing literature on the measurement of inclusive capacity at an individual level. This innovative approach is fundamental, given that there were no previous tools to assess this construct in such a direct and focused manner. It is important to note that the applied methodology ensures face and content validity, conceptual clarity and psychometric coherence of the scale using qualitative methodologies (focus groups, expert consensus, cognitive interviews), as well as the quantitative analysis of the discrimination indexes and psychometric properties of the scale. Additionally, the subsequent score-to-percentile transformation offers an advantage due to its simplicity and correct interpretation. The transformations presented in this paper may be applied to scores of populations comparable to the validation sample, i.e., mostly women who do not identify with any ethnic group and without disabilities.

### Limitations

Although the ICIS scale offers a new perspective on inclusive capacity, it has limitations. The main limitation of this scale is its novelty, which, whilst still being a strength, also implies that it needs to undergo further validation testing and adjustments. On the other hand, our sample was not randomly selected, therefore, it is not representative of the general population. The lack of representativeness and the recruitment process of participants resulted in an overrepresentation of women and other demographic groups, which could compromise its generalizability. In addition to this, the omega coefficient for the Perceived Behavioral Control domain suggests the need for further item development and testing, given its low reliability. All in all, future research is required to validate the scale in more diverse populations, including international contexts, to explore its predictive validity and longitudinal stability, and to perform the corresponding percentile transformation on the scores if desired.

### Implications

The results obtained provide a basis for future research and theoretical development in the field of inclusion. It is essential that future studies explore the adaptability of the instrument in diverse cultural and social settings, improving its generalizability and the understanding of inclusion as a spectrum.

### Conclusions

The ICIS scale offers a novel and robust measure of inclusion intent as a proxy for inclusion capacity. This scale is a valuable tool for evaluating individual-level inclusion capacities at different application levels, including research, clinical practice, and policymaking or policy application. Considering how inclusion has become an increasingly relevant issue across multiple disciplines, the possibility of measuring it reliably holds significant implications for intervention design, policy evaluation, and social impact assessment.

## Author Contributions

L.M.G.B. (Lina María González-Ballesteros): Conceptualization (Ideas; formulation or evolution of overarching research goals and aims), Methodology (Development or design of methodology; creation of models), Supervision (Oversight and leadership responsibility for the research activity planning and execution), Writing – review & editing (Critical review, commentary, or revision of the manuscript).

C.A.C.R. (Camila Andrea Castellanos Roncancio): Data curation (Management activities to annotate data and maintain research data for initial use and later re-use), Formal analysis (Application of statistical, mathematical, computational, or other formal techniques to analyze or synthesize study data), Writing – review & editing (Critical review, commentary, or revision of the manuscript).

L.E.M.O (Luis Eduardo Mojica Ospina): Writing – review & editing (Critical review, commentary, or revision of the manuscript).

## Acknowledgments

We thank the expert committee members, the participants in the focus groups, and the entire team at Fundación Saldarriaga Concha for their contributions. We are also grateful to the AVISINI team for their support throughout the research process.

## Data availability

The data supporting the findings of this study are available upon reasonable request, in accordance with ethical and institutional guidelines and data protection policies.

## Bibliography

Arcieri, A. A. and DeLucia, L. R. (2022) ‘Development of a Scale of Prejudice toward Bisexual and Transgender Individuals on the Basis of Ambiguity Intolerance’, Journal of Bisexuality, 22(1). doi: 10.1080/15299716.2022.2044959.

Association, A. P. (2020) ‘APA Psychnet’, p. 1. Available at: https://www.apa.org/pubs/databases/psycinfo.

Björkman, T., Svensson, B. and Lundberg, B. (2007) ‘Experiences of stigma among people with severe mental illness. Reliability, acceptability and construct validity of the Swedish versions of two stigma scales measuring devaluation/discrimination and rejection experiences’, Nordic Journal of Psychiatry, 61(5), pp. 332–338. doi: 10.1080/08039480701642961.

Boateng, G. O. et al. (2018) ‘Best Practices for Developing and Validating Scales for Health, Social, and Behavioral Research: A Primer’, Frontiers in Public Health, 6(June), pp. 1–18. doi: 10.3389/fpubh.2018.00149.

Campo-Arias, A., Oviedo, H. C. and Herazo, E. (2014) ‘Escala de experiencias de discriminación: Consistencia y estructura interna en estudiantes de medicina Internal consistency and structure in medical students’, Revista CES Psicología, 7(2), pp. 15–26.

Carrillo Sierra, S. M. et al. (2018) ‘Propiedades psicométricas del Cuestionario de Inclusión Educativa (CIE) en contextos escolares colombianos’, Revista Espacios, 39(23).

Chapple, M. and Murphy, R. (1996) ‘The Nominal Group Technique: Extending the evaluation of students’ teaching and learning experiences’, Assessment and Evaluation in Higher Education, 21(2), pp. 147–160. doi: 10.1080/0260293960210204.

Chin, D. et al. (2020) ‘Racial/ethnic discrimination: Dimensions and relation to mental health symptoms in a marginalized urban American population.’, American Journal of Orthopsychiatry, 90(5), pp. 614–622. doi: 10.1037/ort0000481.

Davis, M. H. (1983) ‘A Mulitdimensional Approach to Individual Differences in Empathy’, Journal of Personality and Social Psychology, 44(1), pp. 113–126. doi: 10.1037/0022-3514.44.1.113.

Etchezahar, E. et al. (2022) ‘Assessment of social justice dimensions in young adults: The contribution of the belief in a just world and social dominance orientation upon its rising’, Frontiers in Psychology, 13. doi: 10.3389/fpsyg.2022.997423.

Fiske, S. T. (2009) ‘From dehumanization and objectification to rehumanization: Neuroimaging studies on the building blocks of empathy’, Annals of the New York Academy of Sciences, 1167, pp. 31–34. doi: 10.1111/j.1749-6632.2009.04544.x.

Forbes, C. E. and Grafman, J. (2010) ‘The role of the human prefrontal cortex in social cognition and moral judgment’, Annual Review of Neuroscience, 33, pp. 299–324. doi: 10.1146/annurev-neuro-060909-153230.

de Freitas Nery, N. N., et al. (2023) ‘Scale of Sexual Prejudice Against Bisexuals: Evidence of Validity’, Psico-USF, 28(2), pp. 333–345. doi: 10.1590/1413-82712023280210.

Haslam, N. and Loughnan, S. (2014) ‘Dehumanization and infrahumanization’, Annual Review of Psychology, 65, pp. 399–423. doi: 10.1146/annurev-psych-010213-115045.

Hazen, A. (1914) ‘Storage to be Provided in Impounding Reservoirs for Municipal Water Supply (with discussion)’, Transactions of the American Society of Civil Engineers, 77, pp. 1539–1669. doi: 10.1061/taceat.0002563.

Inc, J. (2025) Jotform.

Janssen, E. M. et al. (2020) ‘Psychometric properties of the Actively Open-minded Thinking scale’, Thinking Skills and Creativity, 36(100659). doi: 10.1016/j.tsc.2020.100659.

Kenny, A., Bizumic, B. and Griffiths, K. M. (2018) ‘The Prejudice towards People with Mental Illness (PPMI) scale: Structure and validity’, BMC Psychiatry, 18(1), pp. 1–13. doi: 10.1186/s12888-018-1871-z.

Klos, M. C. and Lemos, V. N. (2018) ‘Adaptación y validación de un instrumento para evaluar el constructo compasión’, Revista Evaluar, 18(2), pp. 31–44. doi: 10.35670/1667-4545.v18.n2.20801.

Kwan, V. S. Y. and Fiske, S. T. (2008) ‘Missing links in social cognition: The continuum from nonhuman agents to dehumanized humans’, Social Cognition, 26(2), pp. 125–128. doi: 10.1521/soco.2008.26.2.125.

Larsen, K. S., Reed, M. and Hoffman, S. (1980) ‘Attitudes of heterosexuals toward homosexuality: A likert-type scale and construct validity’, The Journal of Sex Research, 16(3), pp. 245–257. doi: 10.1080/00224498009551081.

Londoño Pérez, C. and Alejo Castañeda, I. E. (2017) Instrumentos usados en Colombia para evaluar la dimensión psicológica del proceso salud-enfermedad. Edited by U. C. de Colombia. Bogotá.

López-Pérez, B., Fernández-Pinto, I. and Abad García, F. J. (2008) TECA[: test de empatía cognitiva y afectiva / Belén López-Pérez, Irene Fernández-Pinto y Francisco José Abad García., Test de empatía cognitiva y afectiva. Madrid: TEA.

López Tello, A. and Moreno Coutiño, A. B. (2019) ‘Escala de Compasión (ECOM) para población mexicana’, Psicología y Salud, 29(1), pp. 25–32. Available at: 10.25009/pys.v29i1.2565.

Martínez-Ramos, N., Cárdenas, L. and Aguirre-Acevedo, D. C. (2022) ‘Colombian Adaptation of the Self-Compassion Scale (SCS)’, Psicothema, 34(4), pp. 621–630. doi: 10.7334/psicothema2022.86.

‘MedCalc® Statistical Software version 23.2.1’ (2025). Ostend, Belgium: MedCalc Software Ltd. Available at: https://www.medcalc.org.

Montoya González, S. and Alzate, J. P. (2023) >Índice multidimensional de inclusión social y productiva para personas con discapacidad. Informe nacional. Bogotá.

Morselli, D. and Passini, S. (2012) Measuring Moral Inclusionϑ: A Validation of the Research paper. doi: 10.12682/lives.2296-1658.2012.14.

Nussbaum, M. C. (2011) Creating Capabilities: The Human Development Approach. Harvard University Press.

Passini, S. and Morselli, D. (2017) ‘Construction and Validation of the Moral Inclusion/Exclusion of Other Groups (MIEG) Scale’, Social Indicators Research, 134(3), pp. 1195–1213. doi: 10.1007/s11205-016-1458-3.

Pérez-Fuentes, M. D. C. et al. (2019) ‘The development and validation of the healthcare professional humanization scale (HUMAS) for nursing’, International Journal of Environmental Research and Public Health, 16, pp. 10–12. doi: 10.3390/ijerph16203999.

Pérez-Sáinz, J. P., Calderón-Umaña, R. and Brenes-Camacho, G. (2016) ‘Exclusión social, violencia y ámbito doméstico. Evidencia y reflexiones desde Centroamérica’, Papeles de Poblacion, 22(87), pp. 9–41.

Pettigrew, T. F. and Meertens, R. W. (1995) ‘Subtle and blatant prejudice in western Europe’, European Journal of Social Psychology, 25(1), pp. 57–75. doi: 10.1002/ejsp.2420250106.

Reed, A., Henry, R. and Mason, W. (1971) ‘Influence of statistical method used on the resulting estimate of normal range’, Clinical Chemistry, 17, pp. 275–284. doi: 10.1093/clinchem/17.4.275.

Reysen, S. and Katzarska-Miller, I. (2013) ‘A model of global citizenship: Antecedents and outcomes’, International Journal of Psychology, 48(5), pp. 858–870. doi: 10.1080/00207594.2012.701749.

Rojas Tejada, A. J., et al. (2011) ‘Prejudiced attitude measurement using The Rasch Rating Scale Model’, Psychological Reports, 109(2), pp. 553–572. doi: 10.2466/07.17.PR0.109.5.553-572.

Sánchez, G., Bella, M. N. and Díaz, L. M. (2013) ¡Basta Ya! Colombia: Memorias de guerra y dignidad., Informe General Grupo de Memoria Histórica. Available at: https://centrodememoriahistorica.gov.co/wp-content/uploads/2021/12/1.-Basta-ya-2021-baja.pdf%0Ahttp://www.centrodememoriahistorica.gov.co/micrositios/informeGeneral/descargas.html.

Schoonjans, F., De Bacquer, D. and Schmid, P. (2011) ‘Estimation of population percentiles’, Epidemiology, 22(5), pp. 750–1. doi: 10.1097/EDE.0b013e318225c1de.

Singer, T. and Lamm, C. (2009) ‘The social neuroscience of empathy’, Annals of the New York Academy of Sciences, 1156, pp. 81–96. doi: 10.1111/j.1749-6632.2009.04418.x.

Smith-Castro, V. and Molina Delgado, M. (2011) Cuadernos Metodológicos. La entrevista cognitiva: guía para su aplicación en la evaluación y mejoramiento de instrumentos de papel y lá. San José, Costa Rica: Universidad de Costa Rica.

Spector, P. E., Bauer, J. A. and Fox, S. (2010) ‘Measurement artifacts in the assessment of counterproductive work behavior and organizational citizenship behavior: Do we know what we think we know?’, Journal of Applied Psychology, 95(4), pp. 781–790. doi: 10.1037/a0019477.

StataCorp (2023) ‘Stata Statistical Software: Release 18’. College Station, TX: StataCorp LLC.

Steenbergen, M. (1996) Compassion and American Public Opinion: An Analysis of the NES Humanitarianism Scale. Report to the Board of Overseers of the National Election Stufies. Chapel Hill.

Steindl, S. R. et al. (2021) ‘The Compassion Motivation and Action Scales: a self-report measure of compassionate and self-compassionate behaviours’, Australian Psychologist, 56(2), pp. 93–110. doi: 10.1080/00050067.2021.1893110.

Team, R. C. (2022) ‘R: A Language and Environment for Statistical Computing.’ Vienna, Austria: R Foundation for Statistical Computing.

Tengelin, E. et al. (2019) ‘Constructing the Norm-critical awareness scale: A scale for use in educational contexts promoting awareness of prejudice, discrimination, and marginalisation’, Equality, Diversity and Inclusion: An International Journal, 38(6), pp. 652–667. doi: 10.1108/EDI-10-2017-0222.

Valencia Agudelo, G. D. and Cuartas Celis, D. (2009) ‘Exclusión económica y violencia en Colombia, 1990-2008: Una revisión de la literatura’, Perfil de Coyuntura Económica, (14), pp. 113–134.

Wahl, O. F. (1999) ‘Mental health consumers’ experience of stigma’, Schizophrenia Bulletin, 25(3), pp. 467–478. Available at: https://www.lib.uwo.ca/cgi-bin/ezpauthn.cgi?url= http://search.proquest.com/docview/223547010?accountid=15115%5Cn http://vr2pk9sx9w.search.serialssolutions.com/?ctx_ver=Z39.88-2004&ctx_enc=info:ofi/enc:UTF-8&rfr_id=info:sid/ProQ%3Anursing&rft_val_fmt=info:o.

Waytz, A., Epley, N. and Cacioppo, J. T. (2010) ‘Social cognition unbound: Insights into anthropomorphism and dehumanization’, Current Directions in Psychological Science, 19(1), pp. 58–62. doi: 10.1177/0963721409359302.

Yildirm, C. and Türkglu, A. (2017) ‘Democratic Citizenship Attitude Scale: A Validity and Reliability Study’, Cukurova University Faculty of Education Journal, 46(2), pp. 649–664. doi: 10.14812/cuefd.303672.

